# Benefit and risk associated with interleukin-6 receptor inhibitor administration during severe COVID-19: a retrospective multicentric study

**DOI:** 10.1101/2025.03.04.25323313

**Authors:** Charlène Lefèvre, Théo Funck-Brentano, Marine Cachanado, Alexia Plocque, Audrey Fels, Frederic Pène, Laurent Savale, David Montani, Olivier Voisin, Flore Bintein, Lucille Wildenberg, Axel Philippe, Stephane Legriel, Nicolas Roche, Pierre-Régis Burgel, Marc Tran, Christophe Baillard, Jacques Duranteau, Gilles Chatellier, Francois Philippart

## Abstract

**Background:** During severe and critical COVID-19, therapeutic options remain scarce. Among interventions, the use of interleukin-6 receptor inhibitor (IL-6Ri) is especially controversial due to persistent uncertainty about their efficacy and safety.

**Methods:** We conducted a multicentric retrospective French observational study. All severe or critical COVID-19 requiring hospital admission were included from march 1^st^ 2020 to December 31^th^ 2021. Our main aim was to compare the occurrence of secondary infections function of the administration of IL-6Ri. Digestive, hematological complications and survival were also analyzed.

**Results:** Among 2587 patients requiring hospital admission, 1603 had a severe COVID-19 and 984 a critical one requiring ICU admission. 224 received at least one dose of tocilizumab or sarilumab. Incidence of secondary infection was 29.5% in the IL-6Ri group *vs.* 19.5% without IL-6Ri (unadjusted OR: 1.73 [1.27;2.34]; p = 0.0004) in the whole population. This result remained consistent after adjustment, without multiple imputation (MI) (adjusted OR: 2.12 [1.51; 2.97]; p < 0.0001) and after MI (adjusted OR: 1.47 [1.25; 1.72]; p < 0.0001)). Incidence of hematological or digestive complication were similar between groups. Mortality of patients admitted in ward was higher in the IL-6Ri group (18.7% *vs* 10.5%, p = 0.0155). No difference in 28 days, ICU, hospital of 90 days mortality was noticed among ICU patients.

**Conclusion:** in this population, administration of IL-6Ri was associated with a higher risk of secondary infection in the whole population and with a higher mortality among patients who spent their whole stay in ward.

## Introduction

Since the onset of Coronavirus disease 2019 (COVID-19), numerous therapeutic interventions have been proposed, particularly for severe manifestations of the disease. The efficacy of corticosteroids in severe and critical COVID-19 cases has been swiftly established (1–3). Concurrently, the pronounced inflammatory response observed in severe cases (4) has prompted exploration of cytokine pathway inhibitors as potential therapeutic targets.

Elevated levels of circulating interleukin-6 (IL-6) in critically ill patients, and its correlation with severe hypoxemia, have highlighted the IL-6 pathway as a promising target for intervention (5,6). However, randomized trials investigating IL-6 receptor inhibitors have yielded conflicting results, leading to ongoing uncertainty about their clinical relevance (7), despite numerous meta-analyses (7–10). Moreover, the safety of targeting the IL-6 pathway in septic patients remains to be specifically investigated.

IL-6 receptor (IL-6R) antagonists have been extensively used in chronic inflammatory diseases such as rheumatoid arthritis, juvenile idiopathic arthritis, and Castleman’s disease (11,12). Long-term experience with IL-6 pathway inhibition has highlighted an increased risk of infectious diseases, estimated at approximately 5.5 events per 100 patient-years (7,11). Additionally, altering the IL-6 pathway may compromise digestive mucosa regeneration, mediated by signal transduction through the gp130 protein (12,13). This could potentially lead to upper digestive tract hemorrhage, especially when combined with systemic corticosteroids, and lower digestive tract translocation, contributing to a sustained inflammatory state.

Conversely, the potential risks associated with the acute administration of IL-6 inhibitors are not yet well-documented. This is particularly true in cases of cytokine release syndrome following chimeric antigen receptor T-cell (CAR-T) therapy, or in severe and critical COVID-19 cases (7,14,15). While recent meta-analyses of therapeutic trials have not reported an increased risk of infections (16–18), this remains a topic of debate. Notably, initial studies were primarily designed to assess efficacy rather than to analyze adverse events.

Given the uncertainty surrounding both the efficacy and potential risks, we focused on the safety of IL-6 pathway inhibition in severe and critical COVID-19 cases. The primary aim of our study was to highlight the potential complications associated with the IL-6 receptor inhibitors in use at the time, specifically tocilizumab and sarilumab.

## Methods

### Study design

We conducted a retrospective, multicenter cohort study involving hospitalized patients who had been admitted to eight medical (internal medicine and pulmonary departments) and critical care wards of three regional university hospitals in Paris from March 1, 2020, to December 31, 2021. All consecutive adult patients admitted to these centers during the study period were included if they had documented severe COVID-19, as defined by the IDSA guidelines (19), requiring hospital admission either to a medical ward or an intensive care unit (ICU).

We compared the disease progression between patients who received an anti-interleukin-6 receptor inhibitor (sarilumab or tocilizumab) and those who did not. The primary outcomes were survival, length of ICU and hospital stay, and major adverse events.

The study adhered to the STROBE (STrengthening the Reporting of Observational Studies in Epidemiology) guidelines for reporting observational studies.

### Settings and participants

Five medical departments (three internal medicine and two pulmonology departments) and four intensive care units (ICUs) across three regional university centers participated in the study. The cohort comprised all consecutive adult patients (aged ≥ 18) admitted with a respiratory form of COVID-19, requiring at least nasal oxygen to maintain a pulsed oxygen saturation above 94%. COVID-19 was confirmed by a positive SARS-CoV-2 real-time reverse transcriptase-polymerase chain reaction (RT-PCR) assay from nasal or oropharyngeal swabs, or lower respiratory tract samples (in invasively ventilated patients). Patients without laboratory-confirmed COVID-19 were excluded from the study. Variant determination through systematic sequencing was not available during the study period.

Patients were categorized into two groups: the “ward group,” which included patients who spent their entire hospital stay in a ward, and the “ICU group,” which included patients who required ICU admission at any point during their hospital stay. The percentage of lung lesions identified by CT scan was not recorded due to the heterogeneity in CT scan availability and the variable time delay between hospital admission and imaging during the pandemic.

Patients were admitted to the ICU if they required noninvasive or invasive mechanical ventilation or if they experienced any other acute organ failure. Details are provided in the supplementary material.

### Data collection and proceeding

Data were collected using a standardized electronic form from the day of admission until hospital discharge. Major adverse event included nosocomial infections, myelotoxicity and digestive tract hemorrhage. Hospital acquired and ventilator-associated pneumonia were defined according to the European (20,21) and American guidelines (22). Other ICU-acquired infections were diagnosed by the attending physician following established guidelines (23). Primary bacterial bloodstream infection and central line associated bloodstream infection were defined as in the EUROBACT study (24).

Upper digestive tract hemorrhage was confirmed by endoscopic demonstration of an esophageal, gastric or duodenal lesion (25–27). Lower intestinal hemorrhage was defined by the presence of melena or hematochezia and confirmed by colonoscopy (27). Thrombopenia was defined as a platelet count below 150 G/l; neutropenia was defined as a neutrophils count below 1 500 /mm^3^, with severe neutropenia below 500 /mm^3^.

The primary outcome was the proportion of hospital-acquired infectious episodes during the hospital stay. Secondary outcomes included the occurrence of leukopenia, thrombocytopenia, digestive hemorrhage or perforation, as well as the length of stay in the intensive care unit (ICU) and hospital, and the duration of mechanical ventilation. The exposure variable was the administration of tocilizumab or sarilumab during the hospital stay. Details are provided in the supplementary material.

### Ethical considerations

The study was carried out in accordance with the Declaration of Helsinki and French law. The TOCSIN study was approved by our institutional review board (GERM; IRB n°00012157). Patients were informed about the study and their non-opposition to the collection and use of non-identifying data for research purposes was obtained. The processing of patient data complied with the reference methodology 004 of the French Commission Nationale de l’Informatique et des Libertés (French National Data Protection Agency).

### Statistical analysis

Patient characteristics were described for the entire population and stratified by the use of IL-6 inhibitors (tocilizumab or sarilumab). Categorical variables are presented as counts (percentages), and continuous variables as mean (± standard deviation) or median with interquartile range [Q1; Q3], depending on their distribution. Categorical variables were compared between groups using the Pearson Chi-squared test or Fisher’s exact test, while continuous variables were compared using the Student’s t-test or the Wilcoxon rank-sum test. As there were no formal recommendations for the use of tocilizumab or sarilumab, their prescription was based on patient-specific characteristics and the judgment of the attending physician. To address potential bias introduced by these prescription methods, a logistic regression model was employed to examine the relationship between the use of these medications and hospital-acquired infectious episodes. Details are provided in the supplementary material.

## Results

### Participants

A total of 2587 patients were included in the study over a 22-month period. Of these, 1603 were admitted to a medical ward and remained there for the duration of their stay (referred to as the “Ward group”), while 984 required an ICU admission at some point during their hospital stay. The mean age was 63.6 and 62.7% were male (Table n°1 and supplementary Table 1 and 2).

### COVID-19 severity

High-flow nasal oxygen was required in 81 (10.9%) cases, non-invasive ventilation in 122 (12.5% of ICU patients) and invasive mechanical ventilation in 565 (57.5% of ICU patients) (Table 2 and 3, and supplementary table 3-5).

**Table 1:**
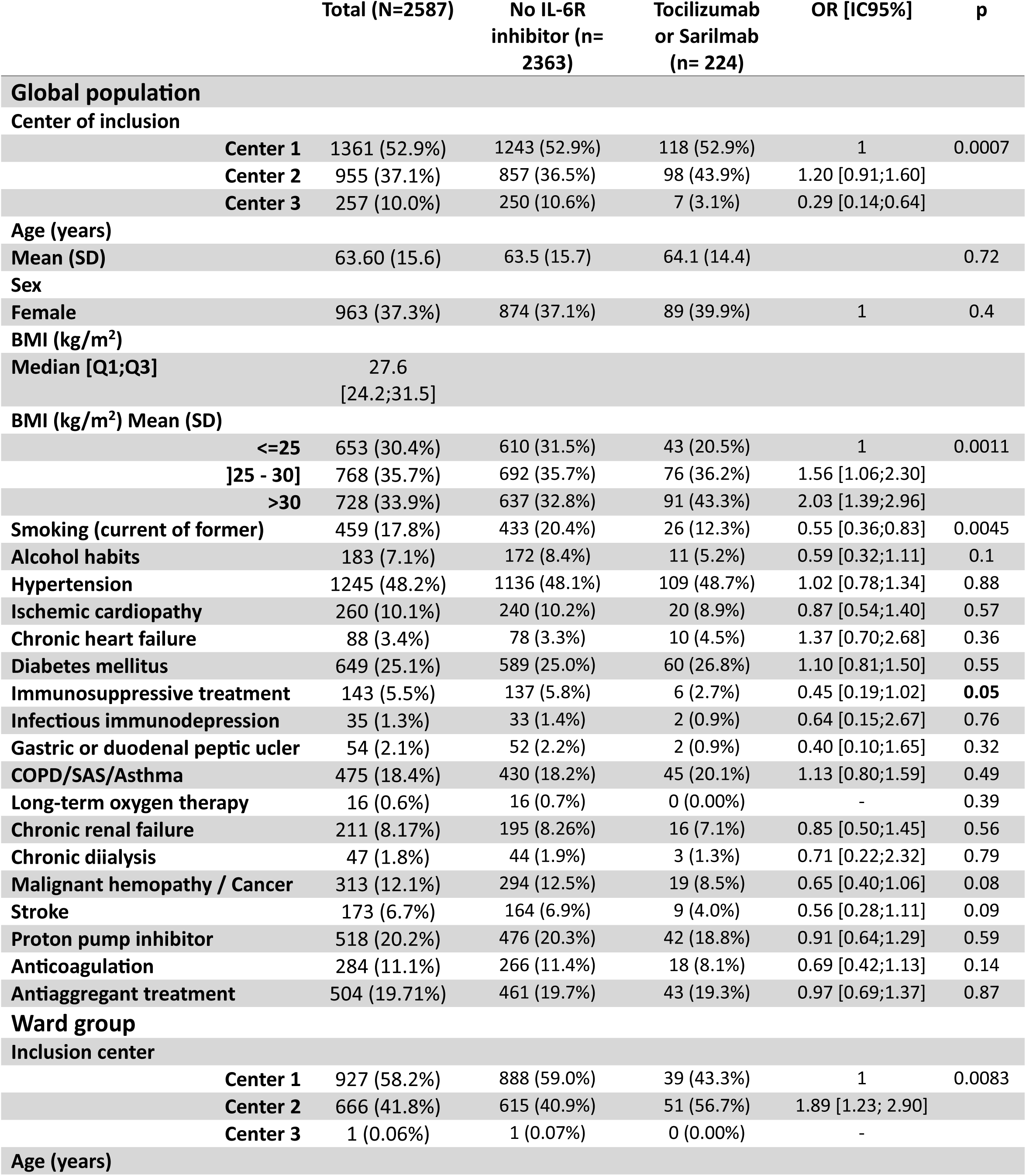
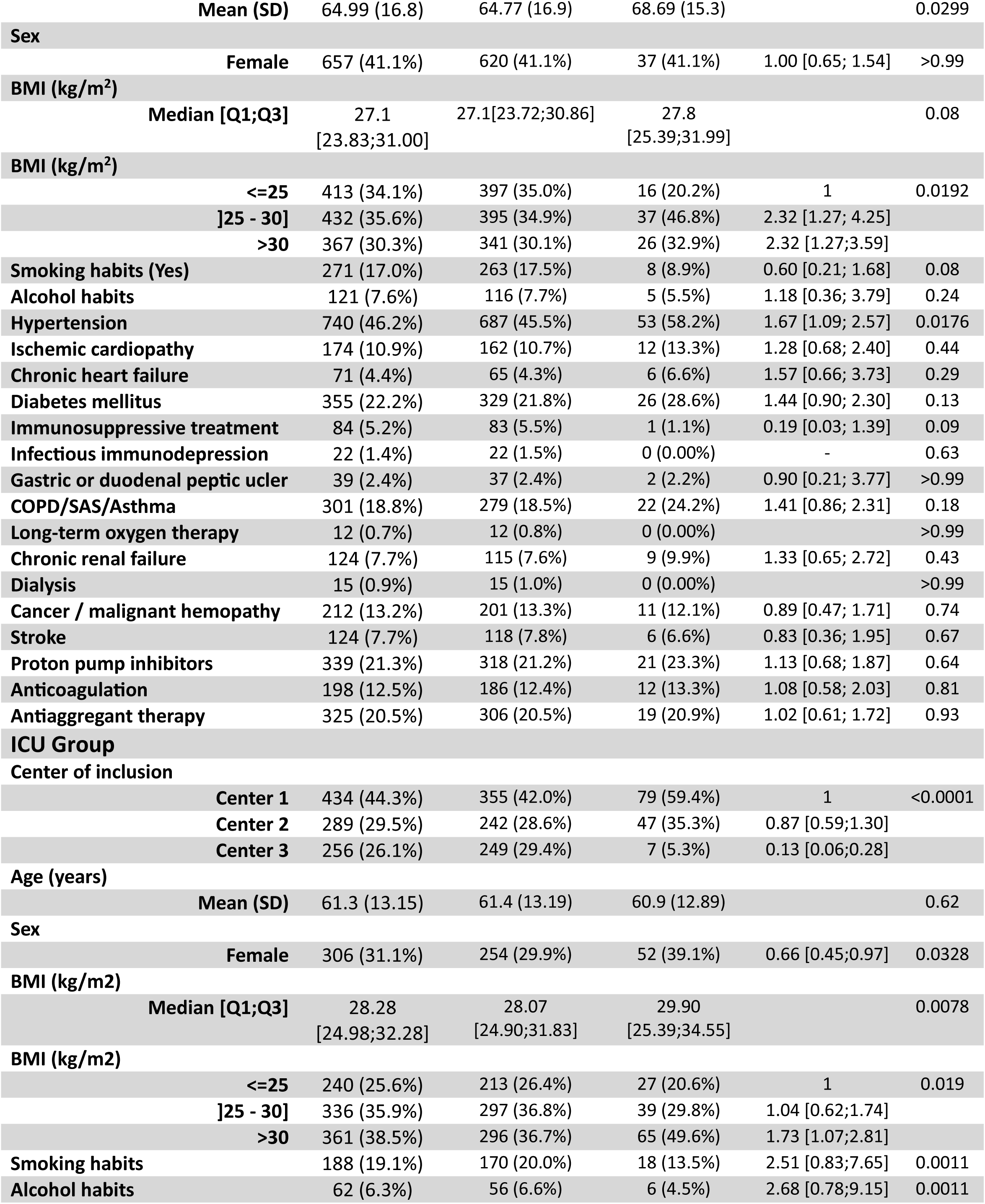
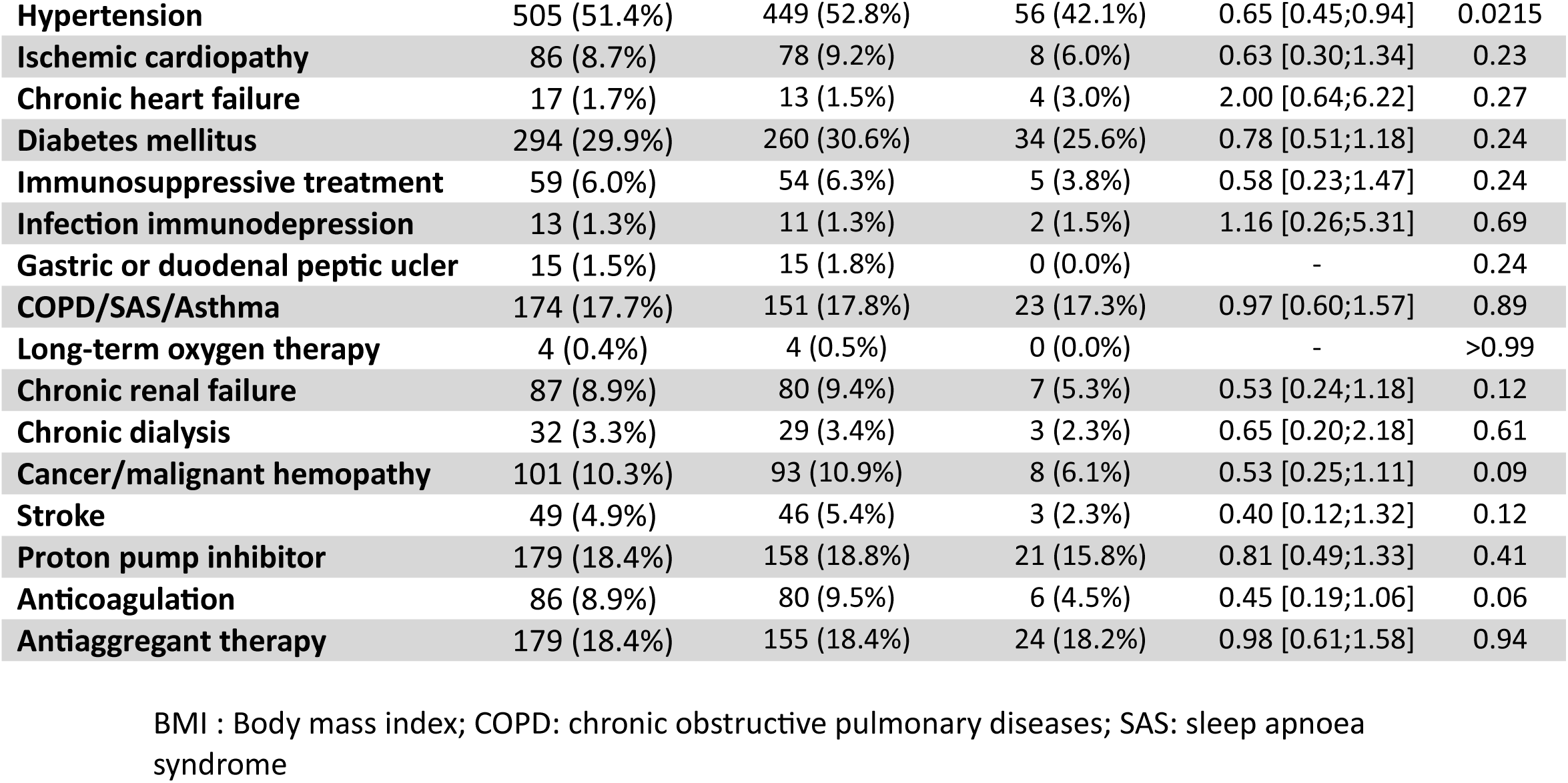
Characteristics of the population.

**Table 2:** Evolution during hospital stay: Ward group.

|  | Total (N=1603) | No IL-6R inhibitor (N=1512) | Tocilizumab or Sarilumab (N=91) | OR [IC95%] | p |
| --- | --- | --- | --- | --- | --- |
| **Type of IL-6R inhibitor** |  |  |  |  |  |
| Sarilumab | 11 (0.7%) | 0 (0.00%) | 11 (12.1%) | - | - |
| Tocilizumab | 80 (5.0%) | 0 (0.00%) | 80 (87.9%) | - | - |
| New anticoagulation | 1461 (91.5%) | 1370 (91.03%) | 91 (100.0%) | 1 | 0.0028 |
| **Anticoagulation. type** |  |  |  |  |  |
| Curative | 367 (25.1%) | 346 (25.26%) | 21 (23.1%) | 1 | <0.0001 |
| Preventive | 652 (44.6%) | 635 (46.35%) | 17 (18.7%) | 0.44<br>[0.23;<br>0.85] |  |
| Intermediate dose preventive | 442 (30.2%) | 389 (28.39%) | 53 (58.2%) | 2.24<br>[1.33;<br>3.80] |  |
| Corticosteroids | 857 (53.5%) | 778 (51.46%) | 79 (86.8%) | 6.21<br>[3.36;<br>11.49] | <0.0001 |
| Proton pump inhibitors | 126 (7.9%) | 97 (6.42%) | 29 (31.9%) | 6.82<br>[4.19;<br>11.10] | <0.0001 |
| Chloroquine/<br>Hydroxychloroquine | 76 (4.7%) | 76 (5.03%) | 0 (0.0%) | - | 0.0195 |
| Anakinra | 58 (3.6%) | 58 (3.85%) | 0 (0.0%) | - | 0.07 |
| Lopinavir/ritonavir | 4 (0.2%) | 4 (0.27%) | 0 (0.0%) |  | >0.99 |
| Standard supplemental oxygen (days) Median [Q1;Q3] | 4.00 [2.00; 8.00] | 4.00 [2.00; 7.00] | 7.50 [5.00; 11.00] |  | <0.0001 |
| High flow nasal oxygen | 122 (7.6%) | 99 (6.58%) | 23 (25.6%) | 4.88<br>[2.91;<br>8.16] | <0.0001 |
| If HFNO. length(days) Median [Q1;Q3] | 5.00 [3.00; 8.00] | 5.00 [2.00; 7.00] | 7.00 [4.00; 10.00] |  | 0.0186 |
| Length of hospital stay (days) Median [Q1;Q3] | 7.00 [4.00; 11.00] | 7.00 [4.00; 11.00] | 10.00 [7.00; 15.00] |  | <0.0001 |

**Table 3:**
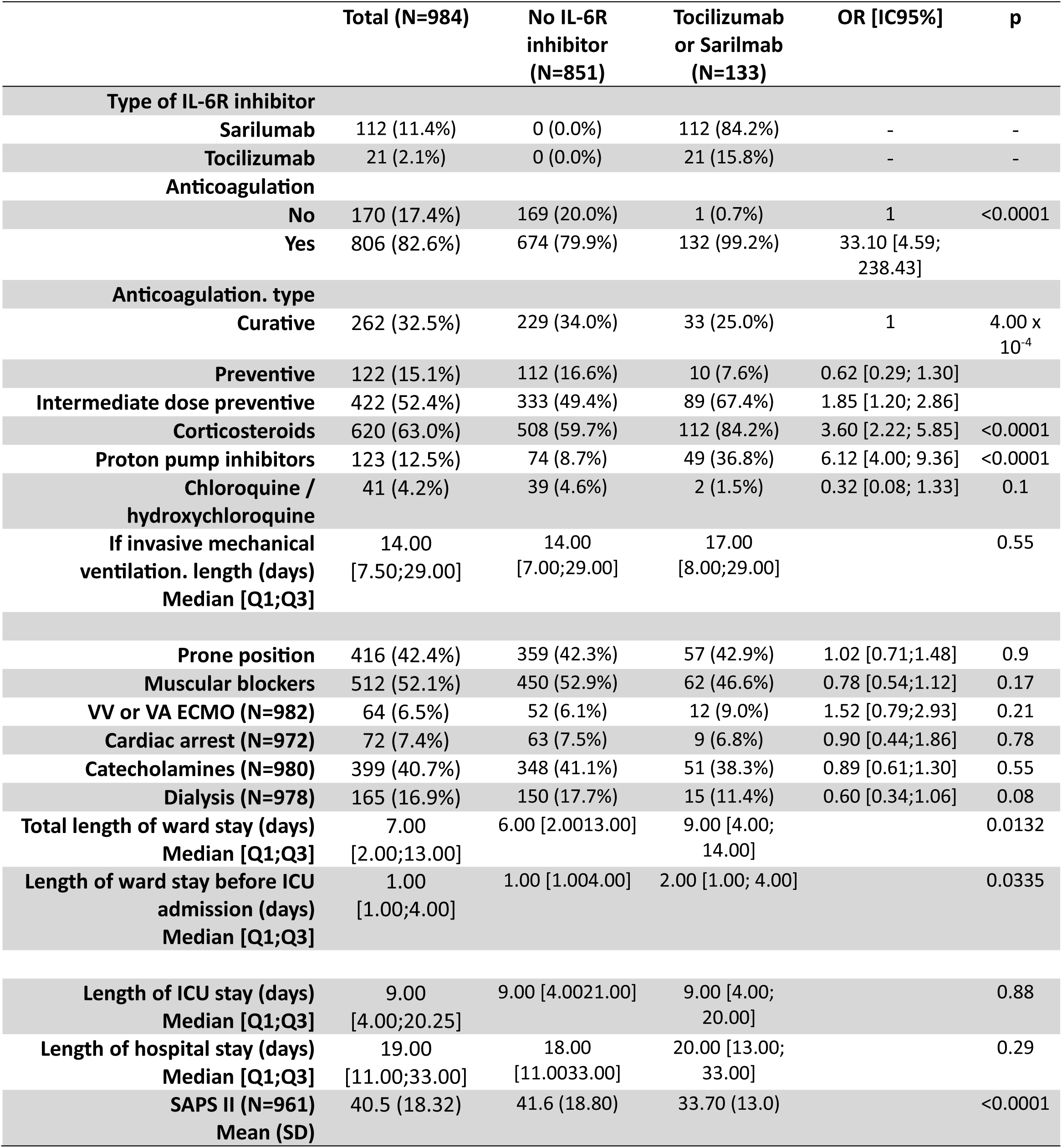
Evolution during hospital stay: ICU Group.

|  | Total (N=984) | No IL-6R inhibitor (N=851) | Tocilizumab or Sarilumab (N=133) | OR [IC95%] | p |
| --- | --- | --- | --- | --- | --- |
| **Type of IL-6R inhibitor** |  |  |  |  |  |
| Sarilumab | 112 (11.4%) | 0 (0.0%) | 112 (84.2%) | - | - |
| Tocilizumab | 21 (2.1%) | 0 (0.0%) | 21 (15.8%) | - | - |
| **Anticoagulation** |  |  |  |  |  |
| No | 170 (17.4%) | 169 (20.0%) | 1 (0.7%) | 1 | <0.0001 |
| Yes | 806 (82.6%) | 674 (79.9%) | 132 (99.2%) | 33.10 [4.59; 238.43] |  |
| **Anticoagulation. type** |  |  |  |  |  |
| Curative | 262 (32.5%) | 229 (34.0%) | 33 (25.0%) | 1 | 4.00 x 10<sup>-4</sup> |
| Preventive | 122 (15.1%) | 112 (16.6%) | 10 (7.6%) | 0.62 [0.29; 1.30] |  |
| Intermediate dose preventive | 422 (52.4%) | 333 (49.4%) | 89 (67.4%) | 1.85 [1.20; 2.86] |  |
| Corticosteroids | 620 (63.0%) | 508 (59.7%) | 112 (84.2%) | 3.60 [2.22; 5.85] | <0.0001 |
| Proton pump inhibitors | 123 (12.5%) | 74 (8.7%) | 49 (36.8%) | 6.12 [4.00; 9.36] | <0.0001 |
| Chloroquine / hydroxychloroquine | 41 (4.2%) | 39 (4.6%) | 2 (1.5%) | 0.32 [0.08; 1.33] | 0.1 |
| If invasive mechanical ventilation. length (days) Median [Q1;Q3] | 14.00 [7.50;29.00] | 14.00 [7.00;29.00] | 17.00 [8.00;29.00] |  | 0.55 |
| Prone position | 416 (42.4%) | 359 (42.3%) | 57 (42.9%) | 1.02 [0.71;1.48] | 0.9 |
| Muscular blockers | 512 (52.1%) | 450 (52.9%) | 62 (46.6%) | 0.78 [0.54;1.12] | 0.17 |
| VV or VA ECMO (N=982) | 64 (6.5%) | 52 (6.1%) | 12 (9.0%) | 1.52 [0.79;2.93] | 0.21 |
| Cardiac arrest (N=972) | 72 (7.4%) | 63 (7.5%) | 9 (6.8%) | 0.90 [0.44;1.86] | 0.78 |
| Catecholamines (N=980) | 399 (40.7%) | 348 (41.1%) | 51 (38.3%) | 0.89 [0.61;1.30] | 0.55 |
| Dialysis (N=978) | 165 (16.9%) | 150 (17.7%) | 15 (11.4%) | 0.60 [0.34;1.06] | 0.08 |
| Total length of ward stay (days) Median [Q1;Q3] | 7.00 [2.00;13.00] | 6.00 [2.00;13.00] | 9.00 [4.00; 14.00] |  | 0.0132 |
| Length of ward stay before ICU admission (days) Median [Q1;Q3] | 1.00 [1.00;4.00] | 1.00 [1.00;4.00] | 2.00 [1.00; 4.00] |  | 0.0335 |
| Length of ICU stay (days) Median [Q1;Q3] | 9.00 [4.00;20.25] | 9.00 [4.00;21.00] | 9.00 [4.00; 20.00] |  | 0.88 |
| Length of hospital stay (days) Median [Q1;Q3] | 19.00 [11.00;33.00] | 18.00 [11.00;33.00] | 20.00 [13.00; 33.00] |  | 0.29 |
| SAPS II (N=961) Mean (SD) | 40.5 (18.32) | 41.6 (18.80) | 33.70 (13.0) |  | <0.0001 |

Median hospital length of stay (LOS) was 7.0 [4.0; 11.0] days for the ward group and 19.0 [11.0; 33.0] days for the ICU group. Among ICU patients, the median time from hospital admission to ICU admission was 1.0 day [IQR 1.0-4.0], and the median ICU LOS was 9.0 [IQR 4.0; 20.3] days (Table 3, Supplementary Tables 3-5).

In the ward group, the median duration of standard oxygen therapy was 4.0 days [IQR 2.0-8.0], and of High-flow nasal oxygen was 5.0 days [IQR 3.0; 8.0] (Table 2, Supplementary Table 4). Median duration of invasive mechanical ventilation was 14.0 days [7.5; 29.0]. Neuromuscular blockade was administered to 52.1% (n=512) of patients, prone positioning was required in 42.4% (n=416) and ECMO support was required in 6.5% (n=64) of patients (Table 3, Supplementary Table 6)

#### COVID-19 Specific Therapeutic Interventions

Corticosteroids were prescribed in 857 patients (53.5%) in the ward group and 620 patients (63.0%) in the ICU group. Other treatments are detailed in Table 3 and supplementary Table 5. With the exception of corticosteroids, COVID-19-specific treatments were initiated during the patient’s ward stay.

A total of 224 patients (8.7%) received at least one dose of tocilizumab or sarilumab. Tocilizumab was administered to 192 patients (7.4%) and sarilumab to 32 patients (1.2%) (Table 2 and 3, Figure 1, Supplementary Tables 1-5). Both IL-6R inhibitors were initiated prior to ICU transfer in the majority of cases (9 out of 133 cases (6.7%) during ICU stay).

**Figure 1:**
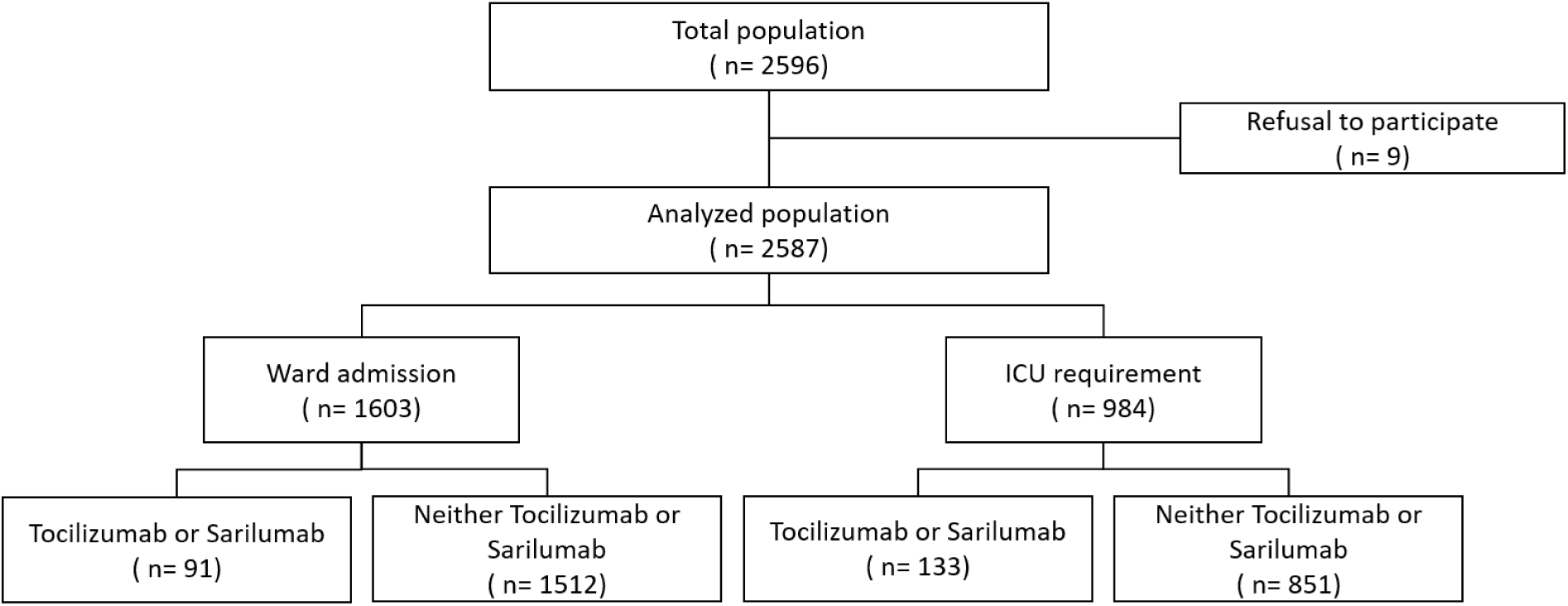
Flowchart.

Patients treated with IL-6R inhibitors received significantly more corticosteroids (85.3% vs. 54.4%, p < 0.0001), proton pump inhibitors (34.8% *vs*. 7.2%, p < 0.0001), and intermediate-dose preventive anticoagulation (63.7% *vs*. 35.3%, p < 0.0001) compared to other patients. These results were consistent in both the ward group (Table 2, Supplementary Table 4) and the ICU group (Table 3, Supplementary Table 5).

#### Outcomes

Among the 2 587 patients included in the study, 2 144 (82.9%) were discharged alive (1427 (89.0%) in the ward group and 717 (72.9%) in the ICU group). In the ward group, mortality was higher in patients receiving IL-6R inhibitors compared to those who did not (17 (18.7%) vs. 159 (10.5%); p < 0.02). In the ICU group, hospital mortality was similar in both groups (31 (23.3%) vs. 236 (27.7%); p = 0.29) (Table 4, Supplementary Table 7).

**Table 4:**
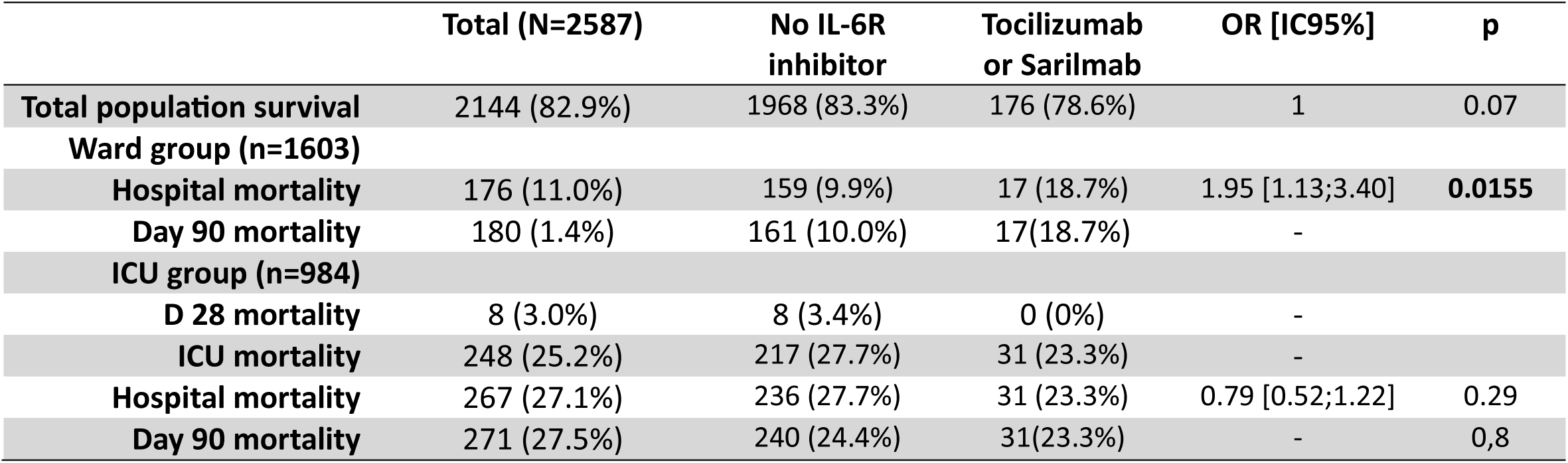
Outcome.

In the ward group, the length of hospital stay was significantly longer in patients receiving IL-6R inhibitors compared to those who did not (10.0 [7.0; 15.0] *vs*. 7.0 [4.0; 11.0]; p < 0.0001) (Table 2, Supplementary Table 4). In the ICU group, no difference in the length of ICU or hospital stay was observed (20.0 [13.0; 33.0] vs. 18.0 [11.0; 33.0]; p = 0.29) (Table 3, Supplementary Table 5).

#### Infectious Complications

During hospital stay, 97 patients (6.1%) in the ward group developed a documented infection. The most frequent were urinary tract infections (53; 3.3%), and pneumonias (6; 0.4%). (Table 5, Supplementary Table 8). Patients who received IL-6R inhibitors developed more hospital-acquired infections than those who did not (unadjusted OR: 1.73 [1.27; 2.34]; p = 0.0004). This result remained consistent after adjustment both with and without multiple imputation (adjusted OR: 2.12 [1.51; 2.97]; p < 0.0001 and adjusted OR: 1.47 [1.25; 1.72]; p < 0.0001, respectively) (Tables 5a, 5b and 6, Supplementary Table 8 and 14). No such difference was observed based on patient severity (medical ward vs. ICU).

**Table 5a:** Clinical course complications: Ward group.

|  | Total (N=1603) | No IL-6R inhibitor (N=1512) | Tocilizumab or Sarilumab (N=91) | OR [IC95%] | p |
| --- | --- | --- | --- | --- | --- |
| **Infection during hospital stay** | 97 (6.0%) | 88 (5.8%) | 9 (9.9%) | 1.78 [0.86;3.65] | 0.11 |
| **Fungal infection** | 2 (0.1%) | 2 (0.1%) | 0 (0.0%) |  | >0.99 |
| **Viral infection** | 1 (0.1%) | 0 (0.0%) | 1 (1.1%) |  | 0.06 |
| **Number of infectious episode (N=95) Mean (SD)** | 1.05 (0.3) | 1.07 (0.3) | 0.89 (0.3) |  | 0.06 |
| **Urinary tract infection (N=1602)** | 53 (3.3%) | 50 (3.3%) | 3 (3.3%) | 1.00 [0.30;3.26] | >0.99 |
| **Primitive bacteremia (N=1600)** | 5 (0.3%) | 5 (0.3%) | 0 (0.0%) |  | >0.99 |
| **Secondary bacteremia (N=1600)** | 10 (0.6%) | 9 (0.6%) | 1 (1.1%) | 1.85 [0.23;14.78] | 0.44 |
| **Catheter related infection (N=1599)** | 1 (0.1%) | 1 (0.1%) | 0 (0.0%) |  | >0.99 |
| **Pneumonia** | 6 (0.4%) | 6 (0.4%) | 0 (0.0%) |  | >0.99 |
| **Any adverse event** |  |  |  |  |  |
| **Number of non-infectious adverse event (Median [Q1;Q3])** |  |  |  |  |  |
| **Hematological complication** | 33 (2.1%) | 29 (1.9%) | 4 (4.4%) | 2.35 [0.81;6.84] | 0.11 |
| - **Thrombopenia (N=32)** | 30 (93.7%) | 26 (92.9%) | 4 (100.0%) |  | >0.99 |
| - **Length of thrombopenia (days) Median [Q1;Q3]** | 2.00 [1.00;4.75] | 2.50 [1.00;5.75] | 1.50 [1.00;2.00] |  | 0.2 |
| - **Neutropenia** | 5 (15.1%) | 5 (17.2%) | 0 (0.0%) |  | >0.99 |
| - **Severe neutropenia** | 1 (3.0%) | 1 (3.4%) | 0 (0.0%) |  | >0.99 |
| **Bleeding** |  |  |  |  |  |
| - **Number of bleeding episodes Mean (SD)** | 12 (0.7%) | 11 (0.7%) | 1 (1.1%) | 1.52 [0.19;11.87] | 0.51 |
| - **Gastroduodenal bleeding** | 1 (8.3%) | 1 (9.1%) | 0 (0.0%) |  |  |
| **Digestive perforation (N=76)** | 1 (8.3%) | 0 (0.0%) | 1 (100.0%) |  | 0.08 |

**Table 5b:**
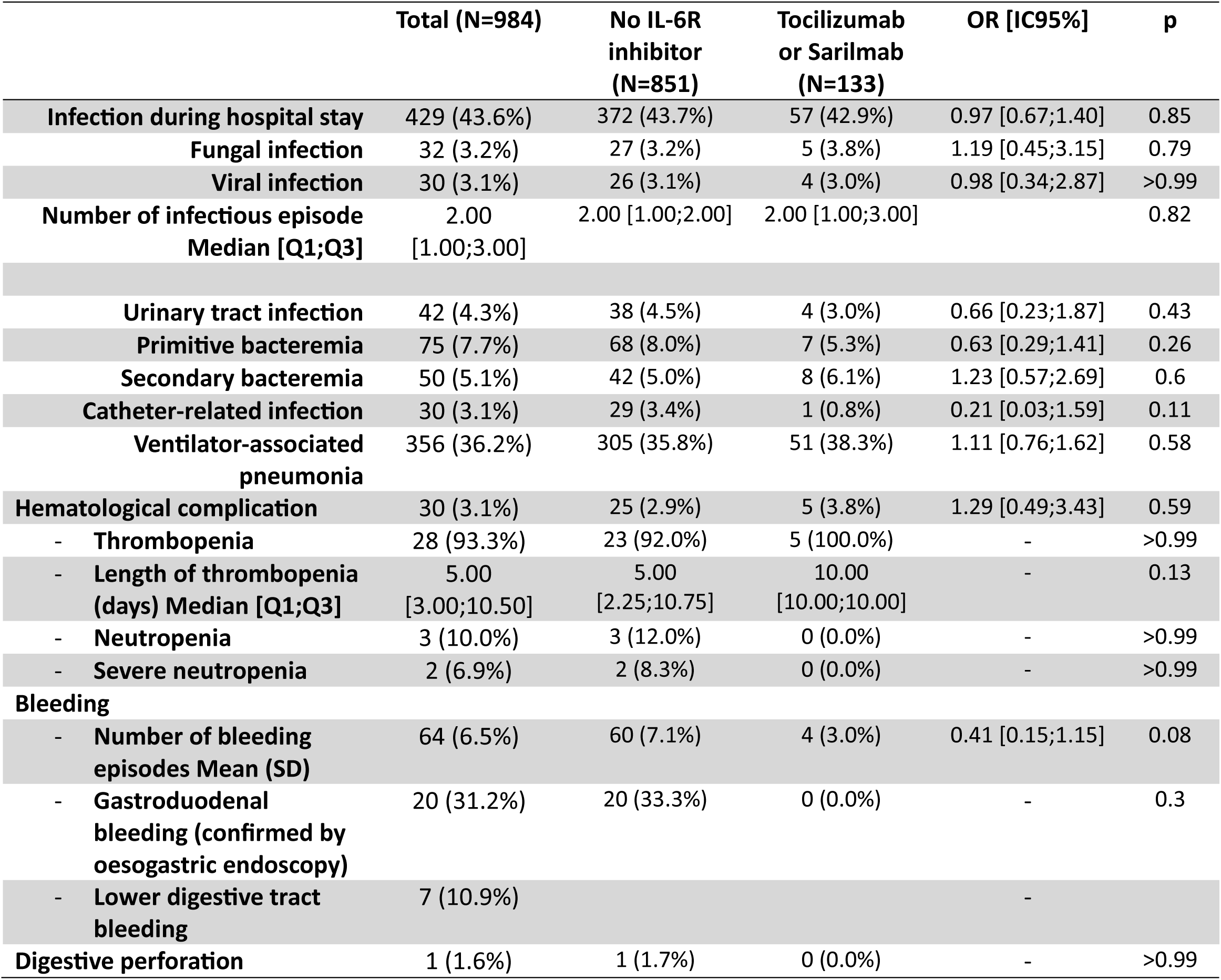
Clinical course complications: ICU group.

**Table 6:**
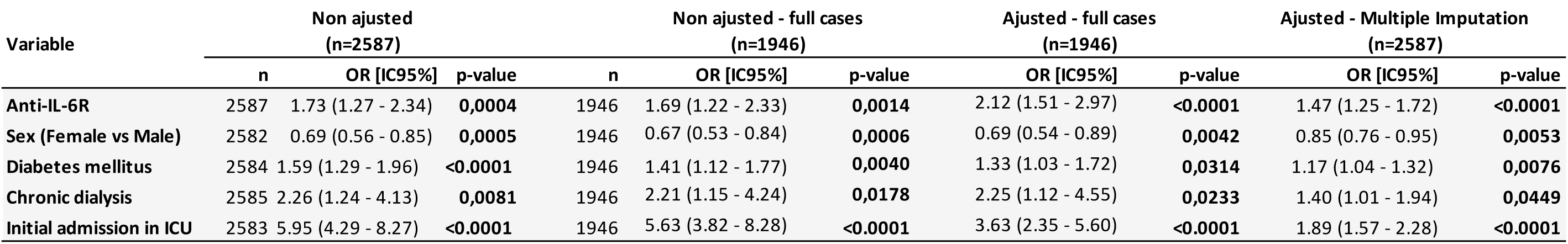
Infectious complication: Multiple imputation.

In the ICU group, the incidence of hospital-acquired infections was similar with or without prior treatment with IL-6R inhibitors (43.7% *vs*. 42.9%; OR: 0.97 [0.67; 1.40]; p = 0.85) (Table 6, Supplementary Table 13).

The combination of IL-6R inhibitors and corticosteroids did not modify the risk of secondary infection compared to corticosteroids alone, neither in the “ward group” nor in the “ICU group” (Table 7). No difference was observed based on the number of initial comorbidities (Supplementary Table 11). However, the risk of infection during ICU stay was associated with the background severity (Supplementary Table 12).

**Table 7:**
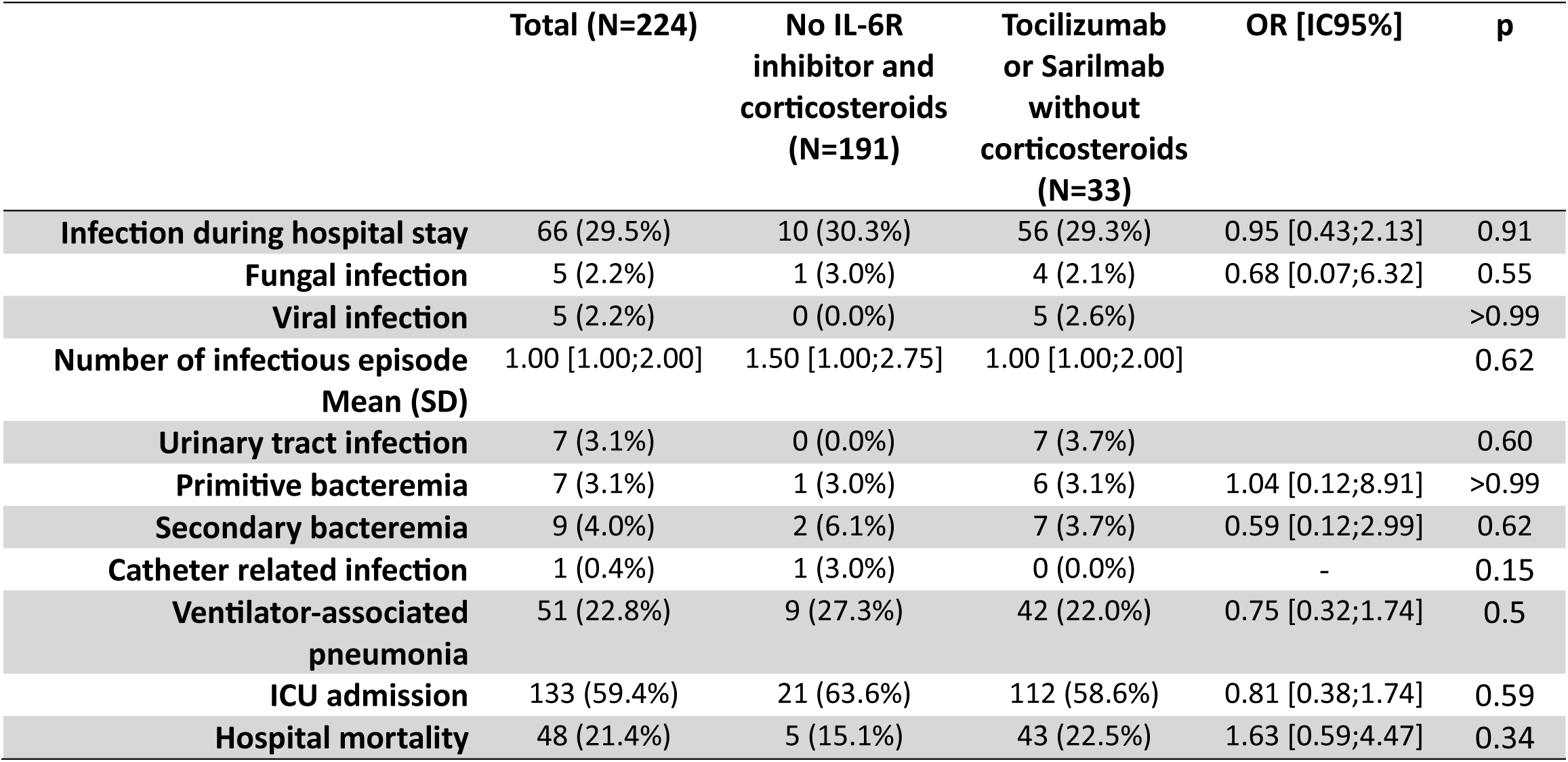
Effect of IL-6R inhibitor and corticosteroids on infection and survival.

#### Other complications

In the ward group, no difference in hematological abnormalities were observed (Table 5, Supplementary Table 10).

A significant digestive tract hemorrhage was noted in one patient (without tocilizumab or sarilumab administration), and a digestive tract perforation occurred in one patient treated with tocilizumab (Table 5).

In the ICU group, hematological complications were noted in 30 patients (3.1%) (Table 6, Supplementary Table 10).

A total of 76 significant hemorrhages (2.9%) occurred in the entire population, with no significant difference based on IL-6R inhibitor administration (Tables 5 and 6).

## Discussion

In this multicenter study, an IL-6R inhibitors were prescribed in 8.6%. The intensity of the inflammatory response associated with SARS-CoV-2 was described early in the pandemic, leading to various attempts to reduce cytokine levels. The efficacy of corticosteroids was first demonstrated (1,2), resulting in their systematic administration. Conversely, the prolonged uncertainty regarding the relevance of IL-6 pathway inhibition (3) led to variable administration during the study period, resulting in a low percentage of patients who actually received an IL-6R inhibitor. This was particularly noticeable in the ICU, where the delayed onset of inflammation and relatively low levels of IL-6 compared to typical acute respiratory distress syndrome or septic shock (4,5) often prevented the administration of these IL-6R inhibitors. As more studies emerged, guidelines were modified based on recent syntheses (6,7). Both suggest the use of tocilizumab in severe COVID-19, but, balance the potential improvements and the prevention of adverse events (7). However, the incidence and severity of these complications remain unclear (3).

We found an increased risk of secondary infections associated with the administration of IL-6R inhibitor in the overall patient population (OR: 1.73 [1.27; 2.34]; p = 0.0004). Impairment of immune defenses due to the disruption of the IL-6 pathway is well-documented during long-term treatment of moderate chronic inflammatory diseases (8,9). Conversely, the effect of a short period of treatment, remains poorly documented. The inability of a single dose to sufficiently dampen the cytokine response seems probable (10) but previous smaller observational studies yielded divergent results. Marco Ripa *et al.* found an increased risk of infections. However, the involvement of IL-6R inhibitors was uncertain, and the multivariate analysis did not confirm this link (11). In another early study in Chicago, the administration of tocilizumab was associated with the risk of secondary bacterial infections (12). Unfortunately, the groups were highly heterogeneous, leading to a disequilibrium that blurred the conclusions. Recent meta-analyses offer similarly conflicting conclusions (13–16).

Many secondary episodes were of viral origin. The central role of IL-6 in viral infection control, by reducing the production of acute phase proteins and modifying B lymphocytes differentiation and T cell maturation, may explain these results (9,17). The absence of observed SARS-CoV-2 clearance compromise during clinical use of tocilizumab (18) tends to reduce the relevance of this hypothesis. However, the timing of use, potential exhaustion of the inflammatory response, and alteration of the respiratory tract mucosa by the initial viral aggression may secondarily favor a new respiratory viral disease. The delay between admission and tocilizumab administration may play a role in the infectious risk (19). Moreover, the association between IL-6R inhibition and specific visceral infections remains unclear. We observed a raw difference in secondary bacteremia that did not reach statistical significance that aligns with previous observations (20,21).

Unexpectedly, while previous studies demonstrated an increased risk of infection in ICU patients (19–21), we did not identify such a difference in our critical care patients. Administration beyond two days of hospital admission or two days of High-flow nasal oxygen requirement has been associated with infectious complications (22). In our study, IL-6R inhibitors were all administered on the first day of hospital admission. This may partly explain the discrepancy in the reported infectious risk between studies and may have contributed to the absence of infections during ICU stay. Conversely, as previously discussed (3), the absence of a deleterious effect is often associated with a lack of beneficial effect in the same studies (3,18). A recent meta-analysis reached a similar conclusion (23). The absence of beneficial effects on survival, organ support requirements during ICU stay, or duration of mechanical ventilation observed in our study may partly explain the lack of deleterious effects from IL-6 pathway inhibition.

Most patients concurrently received corticosteroids, making it challenging to evaluate the separate effects of these drugs (24). The absence of systematic corticosteroid use allowed for an adjustment that confirmed the involvement of tocilizumab, independent of its potential synergistic effect on the immune response (24).

The limited clinical effect of IL-6 pathway inhibitors in our severe and critical population is further supported by the absence of deleterious digestive and hematological effects. Neutropenia has been widely described during tocilizumab or sarilumab treatment, both in chronic inflammatory diseases and COVID-19 (25,26).

Our study has several limitations. First, the retrospective design may result in less comprehensive side effect documentation. However, the severity of the complications of interest makes it unlikely that they were omitted from patient files. Digestive bleeding, perforation, or secondary infections required specific treatment, providing redundant information that made them easier to identify. Furthermore, the incidence of adverse events in our cohort is similar to previous studies and higher than in many randomized trials. Second, the small number of patients treated with IL-6R inhibitors limits the conclusions that can be drawn. This usage however, reflected clinical reality and uncertainty about the benefits and risks of tocilizumab and sarilumab, as indicated by contemporary scientific guidelines. The diversity of wards and ICUs in our study may reflect the heterogeneity of these views, which remain under discussion despite new data and recent meta-analyses. Third, the lack of information on high-flow oxygen use or ICU admission during this critical pandemic period, along with the absence of data on ICU or tracheal intubation withholding, prevents us from drawing conclusions about the potential efficacy of IL-6R inhibition. Nevertheless, adding the results of a cohort study to current and forthcoming high-quality prospective works may have limited relevance.

Conversely, our cohort has several strengths. The multicenter recruitment of severe patients from both wards and ICUs enabled the inclusion of the largest group of patients who received an IL-6R inhibitor in a tolerability study. The retrospective design minimized bias in the reporting of adverse events associated with the administration of controversial therapies. A specific analysis of tocilizumab allowed for the distinction of adverse events associated with the only IL-6R inhibitor shown to have a benefit during severe COVID-19. The multivariate analysis highlighted risk factors associated with an increased probability of secondary infection during IL-6R inhibitor treatment, which should be considered in the risk-benefit analysis of IL-6R inhibitor therapy in this context.

## Conclusion

In this multicenter French cohort study, the administration of an IL-6R inhibitor to severe COVID-19 patients was associated with an increased incidence of secondary infections in ward patients, particularly viral infections. Conversely, in critical cases requiring ICU admission and even invasive mechanical ventilation, no infectious, digestive, or hematological complications were associated with IL-6R inhibitor administration. Broader international studies are needed to confirm these findings.

## Data Availability

All data produced in the present work are contained in the manuscript.

## Acknowledgement

This work was supported by the Groupe Hospitalier Paris Saint-Joseph All data generated or analyzed during this study are included in this study or its supplementary material files. Further inquiries can be directed to the corresponding author.

## Authors contributions

Conceptualization: CL, TFB, AP, MT, FP. Methodology: MC, AF, GC, FP.

Inclusions, review and editing: CL, TFB, AP, FP, LS, DM, OV, FB, LW, AP, SL, NR, PRB, MT, CB, JD.

First draft: CL, TFB, FP.

Conflicts of interest:

Charlène Lefèvre: No conflict

Théo Funck-Brentano: No conflict

Marine Cachanado: No conflict

Alexia Plocque: No conflict

Audrey Fels: No conflict

Frederic Pène: No conflict

Laurent Savale: No conflict

David Montani: No conflict

Olivier Voisin: No conflict

Flore Bintein: No conflict

Lucille Wildenberg: No conflict

Axel Philippe: No conflict

Stephane Legriel: No conflict

Nicolas Roche: Dr Roche reported receiving grant funding from Boehringer Ingelheim, Novartis AG, Pfizer Inc, and GSK PLC and personal fees from Boehringer Ingelheim, Novartis AG, Pfizer Inc, GSK PLC, Austral Pharma, Biosency, MSD, AstraZeneca, Chiesi Farmaceutici SpA, Menarini Group, Nuvaira, Sanofi SA, and Zambon outside the submitted work.

Pierre-Régis Burgel: No conflict

Marc Tran: No conflict

Christophe Baillard: No conflict

Jacques Duranteau: No conflict

Gilles Chatellier: No conflict

Francois Philippart: No conflict

All authors have submitted the ICMJE Form for Disclosure of Potential Conflicts of Interest. Conflicts that the editors consider relevant to the content of the manuscript have been disclosed.

